# Multi-omics-based analysis of the effect of longevity genes on the immune relevance of colorectal cancer

**DOI:** 10.1101/2025.02.13.25322201

**Authors:** Yichu Huang, Guangtao Min, Hongpeng Wang, Lei Jiang

**Author notes:** Correspondence: Jiang Lei. Co-first authors: Yichu Huang and Guangtao Min.

## Abstract

**Background:** Colorectal cancer (CRC) ranks as the third most prevalent cancer globally, with its incidence and recurrence rates steadily rising. To explore the relationship between CRC and longevity-associated genes, and to offer new therapeutic avenues for CRC treatment, we developed a prognostic model based on these genes to predict the outcomes for CRC patients. Additionally, we conducted an immune correlation analysis.

**Methods:** We conducted a comprehensive analysis of the effects of 81 longevity-associated genes in CRC by integrating multiple omics datasets. This analysis led to the identification of two distinct molecular subtypes and revealed that alterations in longevity-related genes across various layers were linked to clinicopathological features, prognosis, and cell infiltration characteristics within the tumor microenvironment (TME). The training and validation cohorts for the models were derived from the TCGA-COAD, TCGA-READ, and GSE35279 datasets. Subsequently, we developed a risk score model, and the Kaplan–Meier method was employed to estimate overall survival (OS). Ultimately, we established a prognostic model based on five longevity-associated genes: BEDN3, EXOC3L2, CDKN2A, IL-13, and CAPN9. Furthermore, we assessed the correlations between the risk score and factors such as immune cell infiltration, microsatellite instability, and the stem cell index.

**Results:** Our comprehensive bioinformatics analysis revealed a strong association between longevity genes and CRC. The risk score derived from the five newly identified longevity-associated genes was determined to be an independent prognostic factor for CRC. Patients categorized by this risk score demonstrated significant differences in immune status and microsatellite instability.

**Conclusion:** Our comprehensive multi-omic analysis of longevity-associated genes highlighted their potential roles in the tumor immune microenvironment, clinicopathological features, and prognosis, offering new insights for the treatment of CRC.

## 1 Introduction

Colorectal cancer (CRC) is the third most common cancer and the second leading cause of cancer death worldwide^1^. Nearly 30% of new CRC cases and deaths worldwide each year occur in China. According to a previous study, the number of newly diagnosed CRC cases in 2022 will exceed 510,000^2^. In addition to previous common treatments, such as surgery, radiotherapy and chemotherapy, in recent years, chimeric antigen receptor (CAR) T-cell treatment^3^, photodynamic therapy^4^, and nanotechnology-based drug/protein delivery systems^5^ have gradually become popular and have shown significant efficacy in antitumor treatment. However, the prevention, diagnosis and treatment of cancer have always been major challenges for scientific research and clinical staff, and improving the survival and health status of elderly patients is still challenging.

As the highest primate of humans, the lifespan of *C. elegans* is no more than tens of days, but humans can live for one hundred years. The longevity genes (anti-aging genes) provide the answer that we want^6^. However, cancer risk should increase with age, which may be related to long-lived organisms having a longer time to accumulate mutations^7^. A longer lifespan is associated with the evolution of more stringent cell cycle control mechanisms, which increase the sensitivity of cells to growth conditions and affect the tumor suppressor effect^8^. A recent univariate analysis for model construction revealed that age, sex, BMI, family history of CRC, smoking status, etc., were independently associated with CRC risk^9^.

Immune senescence is a series of age-related changes that affect the immune system and lead to increased susceptibility to disease over time^10^. As a complex cell network, the tumor microenvironment (TME) is an important factor in regulating the progression of CRC^11^. Immune cells in the TME (cancer-associated fibroblasts, cancer-associated adipocytes, myeloid-derived suppressor cells, etc.) play a role in the development and progression of CRC by producing various cytokines and chemokines and interacting with many chronic inflammatory mediators^12^. The results of immunohistochemistry for mismatch repair proteins (MLH1, PMS2, MSH2, MSH6) and Polymerase Chain Reaction (PCR) for microsatellite instability have prognostic and therapeutic significance in patients with sporadic CRC^13^ and have now become one of the standards guiding clinical medication, but more exploration is needed on individualized treatment options for different types of CRC patients.

In conclusion, immune senescence is recognized for its significant roles in both tumor progression and antitumor responses. A comprehensive understanding of the characteristics of immune infiltration within the longevity-mediated TME may offer valuable insights into the TME of CRC and aid in predicting responses to immunotherapy. These findings pave the way for future research and treatment strategies related to aging, age-related diseases, and longevity.

## 2 Methods

### 2.1 Data download

In this study, we learned from the official The Cancer Genome Atlas (TCGA) website (https://portal.gdc.cancer.gov/repository), which integrates RNA sequencing, genomic mutation data and clinical data (including age, sex and survival status) from the TCGA-COAD and TCGA-READ cohorts. The Gene Expression Omnibus data base (GEO) (https://www.ncbi.nlm.nih.gov) provides mRNA sequencing data and clinical data (GSE35279) from 74 CRC patients, and this dataset underwent quantile normalization and log2 transformation before analysis. In addition, we extracted the copy number variation data of CRC patients from the Xena UCSC database (https://genome.ucsc.edu/). This study was performed in accordance with the Declaration of Helsinki (Revised 2013).

### 2.2 Comprehensive analysis of longevity-related genes in the TCGA database

On the basis of a meta-analysis of individuals from 20 cohorts of European, East Asian or African American descent, gene-level association analysis of tissue-specific gene expression, and a Genome-Wide Association Studies (GWAS) of the strict definition of the longevity phenotype, Joris Deelen^14^ identified longevity-associated genes. The RNA-seq expression data from the TCGA database were first normalized to fragments with a value of millions of bases per thousand, and the RNA-seq expression data from the R language (version: 4.4.1) were first normalized to fragments with a value of millions of bases per thousand^15^. The “limma” package (version 3.56.2) was used to identify differentially expressed genes (DEGs) between normal tissues and tumor tissues; DEGs with screening P values <0.05 are shown in a box plot. The significance level of the DEGs is expressed as follows: * indicates a P value <0.05, ** indicates a P value <0.01, and *** indicates a P value <0.001. Next, the “survival” package of R and the log-rank test^16^ were used. Kaplan‒Meier survival analysis of patients with 34 differentially expressed genes at the intersection of COAD and READ was performed, and the purpose was to compare the survival differences among multiple groups. The mutation data associated with COAD and READ patients in the TCGA dataset were downloaded and processed separately, and the “maftools” R^17^ package was used to illustrate and visualize the most frequently mutated genes. The “Rcircos” package^16^ (version 1.2.2) is helpful for describing the changes in the copy numbers of longevity-related genes on human chromosomes.

### 2.3 Consensus clustering classification

First, RNA expression data and clinical data from the TCGA and GEO databases were integrated using Perl scripts. Next, the “RColorBrewer” package in R was utilized to generate a prognostic network plot of longevity-associated genes. Subsequently, the R package “ConsensusClusterPlus” (version 1.64.0) was applied to the data of all 689 colorectal cancer (CRC) patients from the TCGA and GEO databases to investigate the relationships between significantly expressed longevity-associated genes and CRC subtypes. The optimal K value, representing the clustering variable with the highest correlation within clusters and the lowest correlation between clusters, was selected for classification, and a survival curve was generated.We combined the gene expression and clinical characteristics of all CRC patients in the TCGA and GEO databases, including tumor stage, sex, age, and sample classification, and visualized this data using a heatmap created with the “pheatmap” software package in R. The “limma” package was employed to conduct intercluster variance analysis and principal component analysis (PCA). Based on the PCA results, we identified the common intersection of interacting genes classified as A, C, E, and F to obtain the clustering intersection genes.To facilitate functional enrichment analysis, Weishengxin’s strongest algorithm was used to integrate the Gene Ontology (GO) and Kyoto Encyclopedia of Genes and Genomes (KEGG) databases, allowing for more effective clustering of intersection genes. A significance threshold of P < 0.05 was applied, and results were displayed in histograms and network diagrams. Additionally, the “limma” and “ggpubr” packages in R were used to conduct univariate Cox regression analysis on the cluster intersection genes, enabling the screening and visualization of prognosis-associated genes in a forest plot. Finally, we retyped and screened the samples based on the prognostic genes, selected the most appropriate K value for sample typing, identified differentially expressed genes (DEGs), and generated relevant survival curves.

### 2.4 Construction of the prognostic model

After the CRC prognostic risk genes were incorporated into the model, we used minimum absolute shrinkage and selection operator (LASSO)^18^. The algorithm was employed for dimensionality reduction to identify prognostic feature genes. The risk score was calculated using the following formula: (expression of each signature gene* model coefficient value). Patients were subsequently divided into high-risk and low-risk groups based on the calculated average risk score. The “ggalluvial” package in R was utilized to construct the prognostic models and create the Sankey diagram. To analyze the differences in risk scores between the initial clustering and gene clustering, the “limma” package was employed, and the results were visualized using box plots generated with the “ggpubr” package. Patients were classified into high-risk or low-risk categories according to the median prognostic risk score. The CRC patients were evenly split into a training group and a test group. Kaplan‒Meier survival curves were generated using the “survival” package in R to assess significant differences in overall survival (OS) between these groups. The effectiveness and accuracy of the models were evaluated by measuring the area under the curve (AUC) for 1, 3, and 5 years. The risk curve function was defined in R, and risk curves, survival state maps, and risk heatmaps for both the training and test groups were generated. Additionally, a nomogram model for clinical prediction was constructed by integrating the risk score model with key clinicopathological parameters (sex, age group, tumor stage, risk value, total score, and linear predictive value). “time ROC”^19^ package analysis was used to evaluate the performance and accuracy of the predictive features. In addition, the R “include” function was used to draw mutational waterfall plots of the low-risk and high-risk groups.

### 2.5 Immune, mismatch repair and stem cell index analysis

Analysis of immune cell infiltration in tissues is a key part of disease characterization and prediction of disease prognosis. Using the CIBERSORT algorithm^20^, the relative content of each immune cell in the immune microenvironment of high-risk and low-risk samples was evaluated. All immune cells were then analyzed to construct a scatter plot and a correlation heatmap illustrating the relationship between the risk score and immune cell types. The “estimate” package in R was employed to assess the abundance of stromal and immune cells in each sample, while the “ggplot2” package was used to create violin plots for visualization. The results of the differential analysis among the microsatellite stability group, low-frequency microsatellite instability group, and high-frequency microsatellite instability group were presented as percentage histograms and box plots. Furthermore, the R package “ggExtra” was utilized to calculate the correlation between the risk score and the stem cell index score.

### 2.6 Statistical analysis

To standardize the processing of transcriptomic, clinical, and gene expression data in this study, PERL software (version 5.30.0) was used. All the statistical analyses were performed via R version 4.4.1, and the significance level was set at P <0.05.

## 3 Results

### 3.1 Correlation analysis of longevity-related genes in the TCGA database

DEGs between normal tissues and CRC tissues were analyzed, and these DEGs were compared with longevity-associated genes. A Venn diagram was utilized to identify overlapping DEGs in CRC and rectal cancer (RC) tissues, resulting in 36 common longevity-related genes (Figure 1A). Our analysis revealed that 44 longevity-related genes were upregulated in CRC, while 19 genes were upregulated in RC. The intersection genes identified in the survival analysis for CRC included RAD50, ELOVL6, CDKN2A, MAGI3, BEND3, ZCHC1, DPP4, CELSR2, PSRC1, SNX29, CAPN9, HLA-DRB1, CHRNA3, HLA-DQA1, EPHX2, TOX, and C1orf198. Notably, the expression of KLF6 was found to impact patient survival. The mutation frequencies of these genes were analyzed using a heatmap generated from the TCGA dataset. IGF2R exhibited a high mutation frequency in both colon and rectal cancers, with the majority of mutations being missense mutations (Figure 1C). Copy number variations (CNVs) were observed across multiple chromosomes; however, mutations associated with CRC were predominantly located on chromosome 1 (Figure 1B). Additionally, CNV analysis indicated a significant increase in the proportion of CNVs for CHRNA4 and USP42 in colon cancer, while an increase in CNVs for FURIN and FES was noted in rectal cancer. Conversely, a significant loss of CNVs for LPL was observed in CRC (Figure 1D). Furthermore, the network diagram results confirmed the existence of interactions and tight regulatory connections among longevity-related genes (Figure 2A).

**Figure 1.**
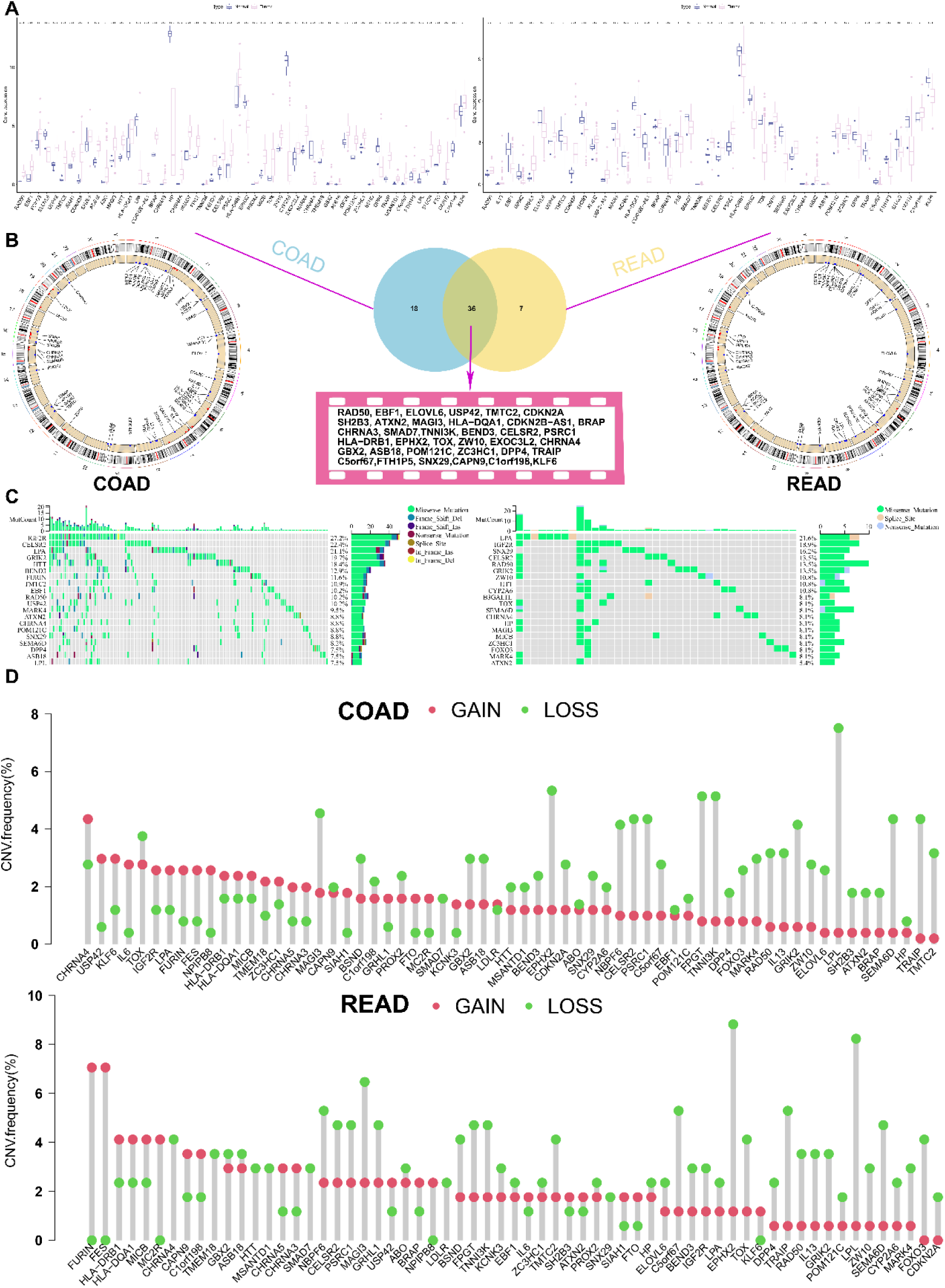
Analysis of cell longevity-related genes in TCGA. (A) Longevity-related genes differences between tumour and normal tissues in the TCGA database, *, P<0.05; **, P < 0.01; ***, P < 0.001. (B) locations of COAD and READ alterations in the longevity-related genes on 23 chromosomes. (C) Waterfall diagram of longevity-related genes mutations. (D) loss and gain of copy number of longevity-related genes; TCGA, The Cancer Genome Atlas; COAD, Colon adenocarcinoma; READ, Rectum adenocarcinoma

**Figure 2.**
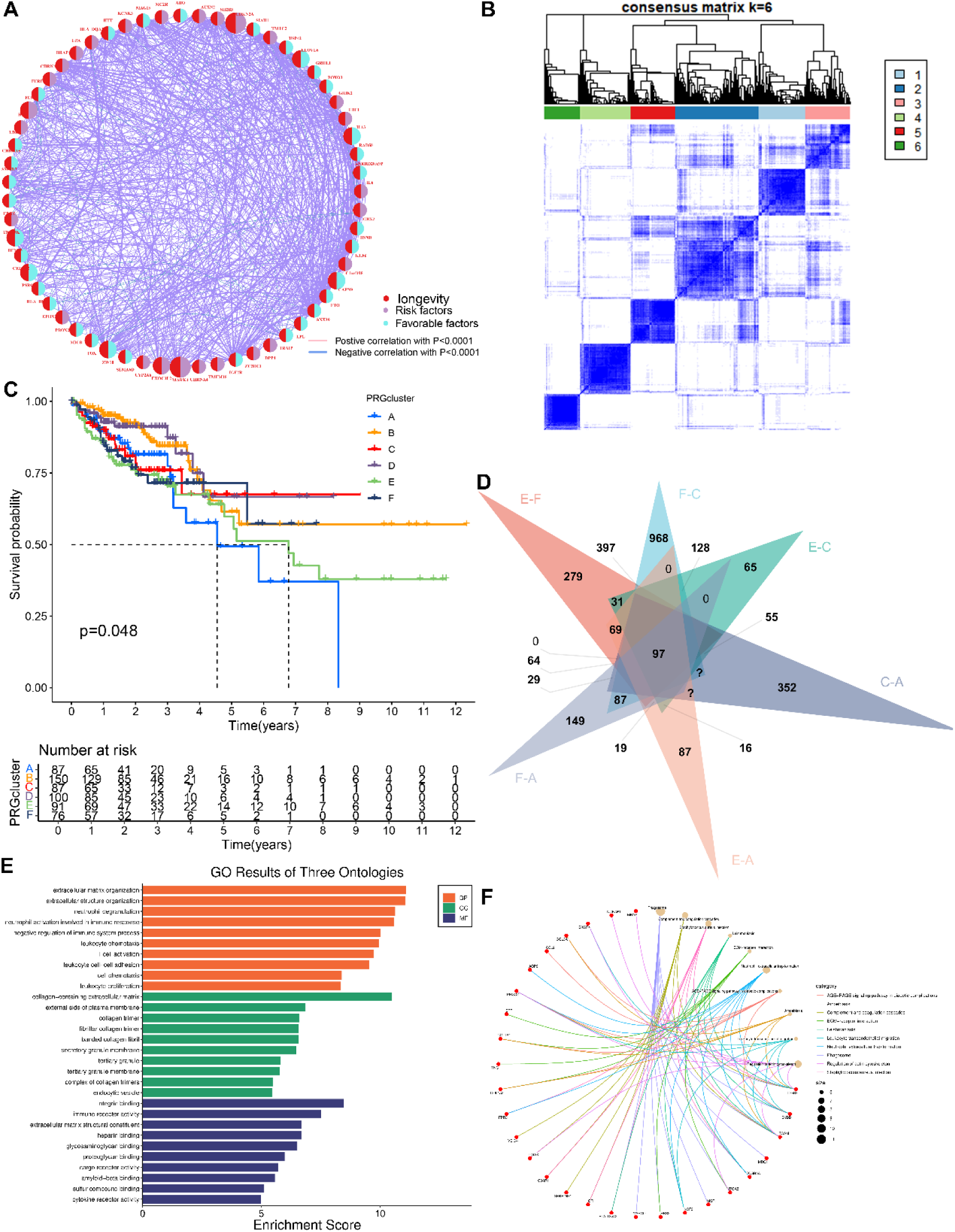
Primary typing of longevity-related genes. (A) Prognostic network of longevity DEGs; (B) a total of 698 patients with CRC were divided into six clusters according to the consensus clustering matrix (k=6); (C) Kaplan-Meier overall survival curves of six clusters; (D) Take the intersecting genes for clusters A, C, E, and F; (E) bar chart of Gene Ontology (GO) pathways; (F) network diagram of Kyoto Encyclopedia of Genes and Genomes (KEGG) pathways.

### 3.2 Construction of the clustering model

To gain a deeper understanding of the expression characteristics of DEGs in CRC, we employed a consensus clustering method. Initially, we integrated data from patients in the TCGA and GEO databases and conducted consistent cluster analysis. When the clustering variable (K=6), we observed the highest homogeneity within groups and the lowest correlation between groups, resulting in the division of the 689 CRC patients into six clusters, each exhibiting significant differences in survival (Figure 2B-C). A comparison of immune cell infiltration across the six clusters revealed varying levels of activated immune T cell enrichment among the clusters. Principal component analysis (PCA) indicated low correlation among Clusters A, C, E, and F (Figure S2A), and significant differences in survival were noted (Figure 2C). The heatmap further illustrates the clinical characteristics of patients within the different clusters (Figure S2C). These 36 DEG-based subtypes effectively differentiated CRC patients, providing crucial evidence for personalized treatment approaches. Additionally, we utilized the Venn diagram function to intersect the differentially expressed genes in patients from Clusters A, C, E, and F, resulting in 97 co-expressed clustered differential genes. GO and KEGG functional enrichment analyses revealed that these genes were primarily associated with neutrophil degranulation, leukocyte chemotaxis, proliferation, differentiation and intercellular adhesion, T cell activation, leukocyte transendothelial migration, chemokine signaling pathways, and the PI3K-Akt signaling pathway (Figure 2E-F). The clinical significance of the 97 clustered intersection genes was further analyzed through univariate Cox regression, identifying 10 genes with notable prognostic value. Specifically, IL-13, ELOVL6, BEND3, PSRC1, ZW10, and CAPN9 were recognized as protective factors, while CDKN2A, FES, EXOC3L2, and MAPK4 were identified as risk factors (Figure 3D). Based on these significant prognostic genes, a second gene cluster analysis was conducted, resulting in the classification of samples into two groups, A and B (Figure 3A). Survival curve analysis demonstrated a significant difference between the two groups (P = 0.006), with patients in group A exhibiting higher survival scores than those in group B. We also analyzed the expression differences of longevity-associated genes between the two genotypes, revealing statistically significant differences in the expression of 38 longevity-associated genes between type A and type B patients (Figure 3C). A Sankey diagram illustrates the classification process (Figure 4A). The heatmap presents gene expression profiles along with clinical characteristics (tumor stage, age, etc.), highlighting a significant age difference among patients across the different groups (Figures S2C-D). From the prognostic genes, five were selected through LASSO regression analysis (Figure S2E-F), leading to the formulation of the risk score calculation: ((IL-13 \times −1.82) + (CDKN2A \times 0.18) + (BEND3 \times −0.50) + (EXOC3L2 \times 0.49) + (CAPN9 \times −0.12)). This formula was utilized to predict risk scores for samples in the training group, which were subsequently divided into high-risk and low-risk categories based on the median risk score. To evaluate the robustness of the risk score, we conducted differential analysis of risk scores between the initial and second genotyping groups, confirming reliable results (Figure 4B-C). Based on the calculated mean risk score, we analyzed the differences in longevity-associated genes between the high- and low-risk groups, identifying 24 longevity-associated genes with significant expression differences (Figure 4D). Furthermore, we examined the mutational landscape of the five prognostic genes in both risk groups, noting that BEND3 exhibited the highest frequency of missense mutations (Figure S2G).

**Figure 3.**
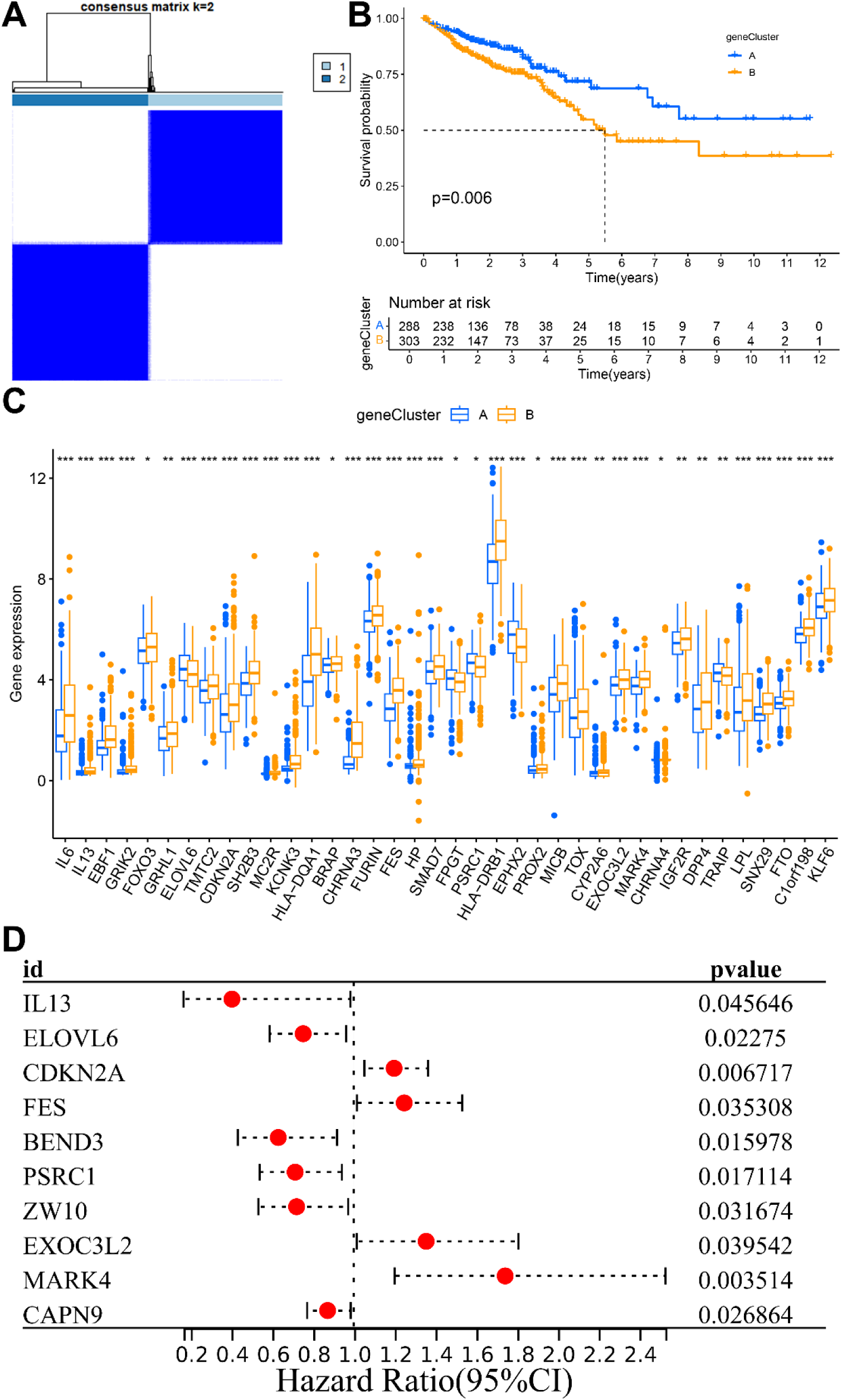
Second typing of cell longevity-related genes (gene clusters). (A) A total of 698 patients with CRC were divided into two clusters according to the consensus clustering matrix (k=2); (B) Kaplan-Meier overall survival curves for two clusters; (C) Box-and-line plot showing differential expression of longevity-related genes in A and B component phenotypes, *, P<0.05; **, P<0.01; ***, P<0.001. (D) Forest plot showing prognosis-related genes

**Figure 4.**
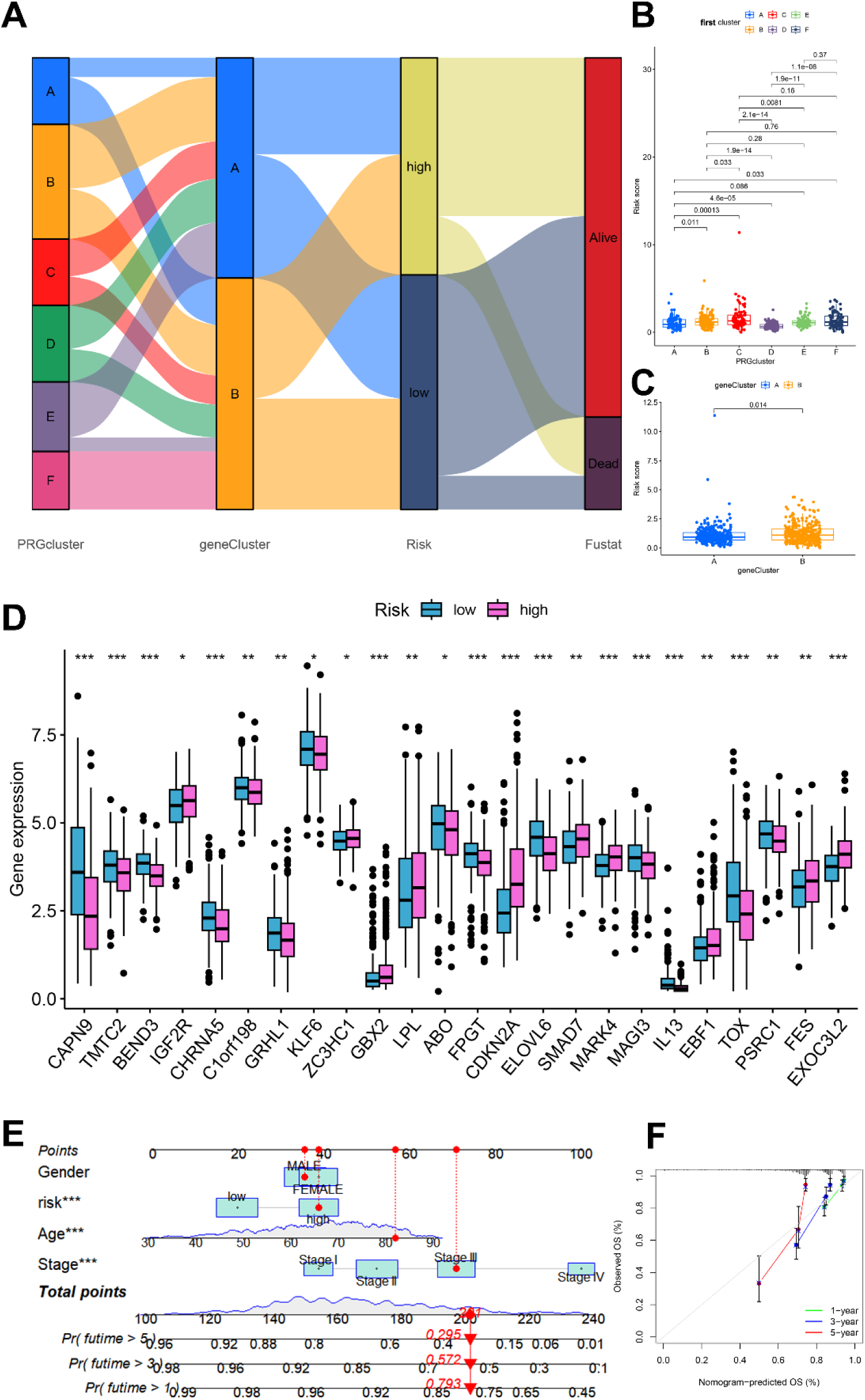
Clinical analysis of the prognostic models. (A) Sanggi diagram of the prognostic model; (B-C) Risk score of the longevity-related genes clusters and gene clusters; (D) longevity-related DEGs in the risk groups; (E) nomograph focusing on factors;(F) predicted 1-, 3-, and 5-year survival calibration curve.

### 3.3 Construction and validation of the clinical prediction models

Based on the prediction model derived from the risk score, we performed univariate and multivariate Cox regression analyses on clinical and pathological features, including patients’ sex, age, and tumor stage, to construct a clinical prediction nomogram model (Figure 4E). Encouragingly, the index curve confirmed the model’s accuracy in predicting survival rates, demonstrating superior performance compared to clinical factors such as age, sex, and stage. This indicates that the model is highly accurate in predicting the 1-, 3-, and 5-year survival rates of patients (Figure 4F). To assess the reliability of the risk model, we conducted a differential analysis of survival between high-risk and low-risk groups. Survival curves indicated that patients in the low-risk group exhibited better survival scores compared to those in the high-risk group, both in the training and test groups, as well as in the overall sample (Figure 5A). The area under the curve (AUC) values for the 1-, 3-, and 5-year survival rates were all greater than 0.5, further confirming the robustness of the results (Figure 5B). Additionally, we categorized patients into high-risk and low-risk groups based on risk scores from the training, test, and overall samples (Figure 5D). An increase in risk score corresponded to a heightened risk of mortality, with the probability of survival gradually decreasing as the duration of survival increased (Figure 5E). The risk heatmap illustrated that the expression of high-risk genes, specifically CDKN2A and EXOC3L2, increased alongside rising risk scores (Figure 5C). These results indicate that the prognostic model can effectively predict the prognosis of CRC patients.

**Figure 5.**
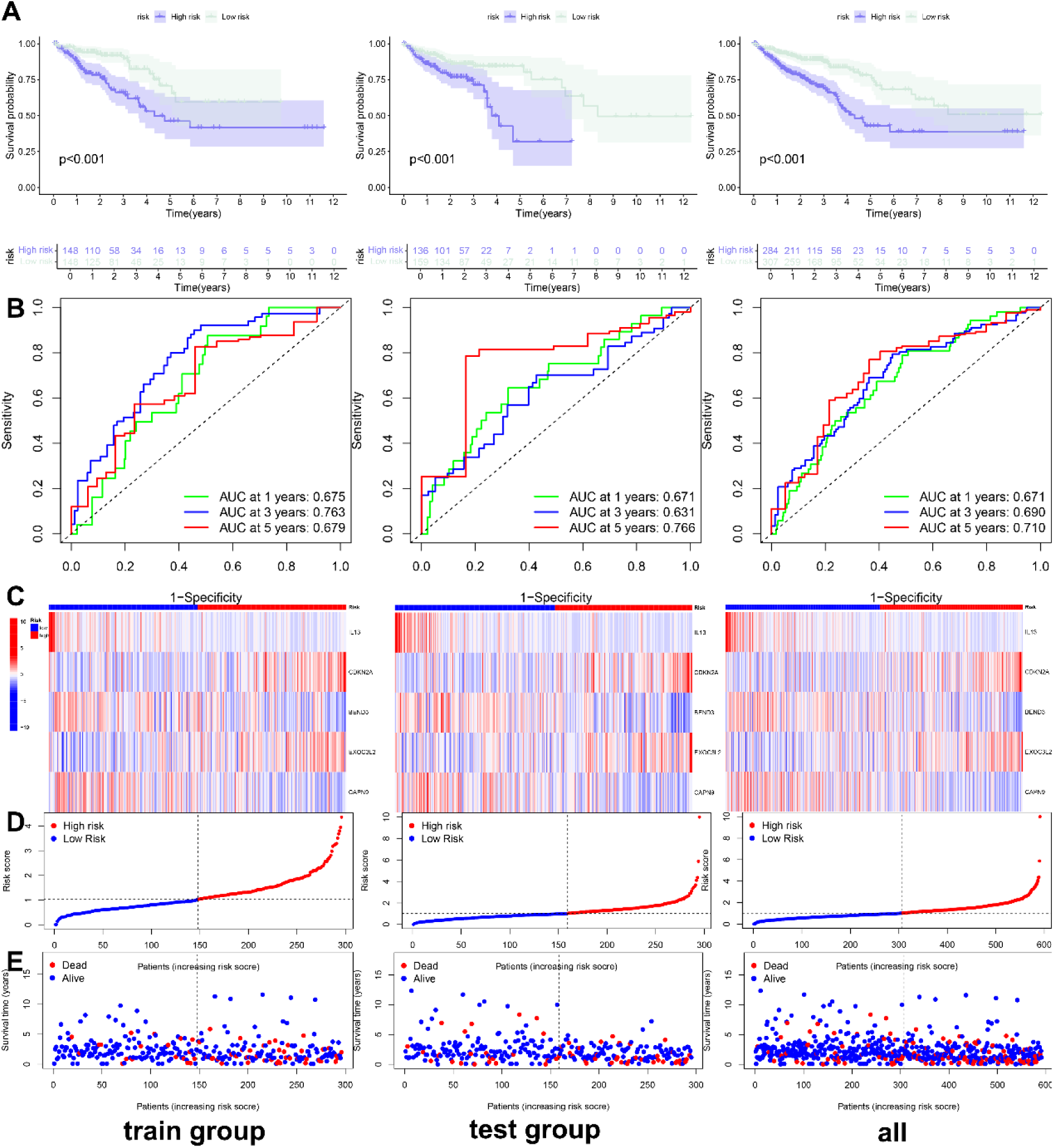
Validation of prognostic model. (A) Survival curve of the training, test and all group; (B) receiver operating characteristic (ROC) curve of the training, test and all group; (C) risk heatmap of the survival-related genes in the training, test and all group; (D) risk curve of the survival-related genes in the training, test and all group; (E) survival state diagram of the survival-related genes in the training, test and all group.

### 3.4 Immune cell and mismatch repair assays

Through the analysis of the five signature prognostic genes, we observed that CAPN9 expression was positively correlated with resting memory CD4 T cells, EXOC3L2+ T cells, regulatory T cells (Tregs), M0 macrophages, IL-3+ T cells, and eosinophils. In contrast, CAPN9 exhibited a significant negative correlation with M1 macrophages, EXOC3L2 macrophages, eosinophils, CDKN2A+ T cells, and resting memory CD4+ T cells (Figure 6A). Notably, higher risk scores were associated with a lower abundance of activated dendritic cells, activated mast cells, resting memory CD4+ T cells, and the infiltration coefficients of neutrophils and eosinophils (Figures 6B-H). These findings suggest that a low risk score is closely related to characteristics of immune cell activation, while a high risk score may correlate with characteristics of matrix activation (Figure 6I). Additionally, the percentage of microsatellite instability-high (MSI-H) patients in the low-risk group was greater compared to those in the high-risk group, and the risk scores for MSI-H patients were lower (Figures 6J-K). Interestingly, a negative correlation was observed between the stem cell index and risk score in CRC patients (Figure 6L).

**Figure 6.**
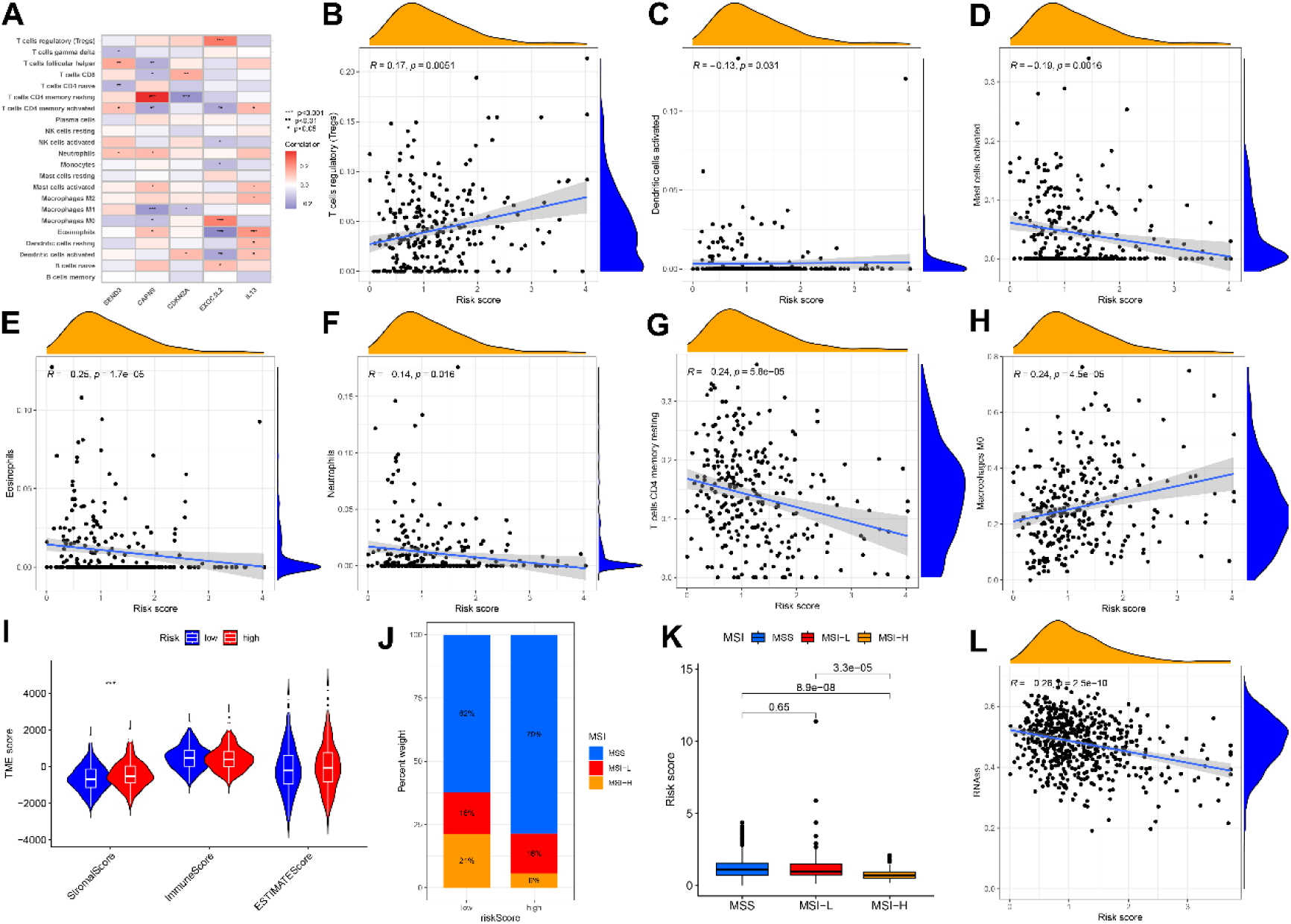
Analysis of the immune microenvironment of the prognostic models. (A) Correlation graph between the risk score and each immune cells; *, P<0.05; **, P<0.01; ***, P<0.001. (B-H) Correlation graph between the risk score and each immune cells; (I) differential analysis of tumor immune microenvironment. (J, K) Relationship between risk scores and microsatellite instability; (L) Scatterplot Demonstrates Association of Risk Score with Tumor Stem Cell Index.

## 4 Discussion

Human longevity is a phenotype that integrates many aspects of health and the environment into one ultimate quantity: the elapsed time from birth to death^21^. The study of the genetic components of human longevity can provide important insights for the prevention of age-related diseases and the mechanisms of various diseases. However, only a limited number of robust longevity genes have been detected in genome-wide association studies^22^. Age is an independent prognostic factor for a variety of malignant tumors, and the activation of longevity genes has been confirmed to have strong tumor-promoting effects. However, cell proliferation arrest induced by progressive telomere shortening (replicative senescence) mediated by telomere genes is also a key human anticancer factor. One of the mechanisms^23^.

The Chinese guidelines for CRC diagnosis and treatment recommend that people after the age of 40 undergo colonoscopy for CRC screening every 5–10 years^24^. However, this will face an enormous cost and resource burden. Therefore, innovative CRC screening indicators, blood-based biomarker detection and stool DNA detection have shown potential for improving the screening participation rate and the sensitivity of precancerous lesions. On the other hand, in response to the heterogeneity of CRC patients, the development of individualized treatment regimens has become a hot topic of current research. Some clinical trials already include combinations of chemotherapy, radiotherapy, hormone therapy and immunotherapy. The combination of cytotoxic compounds and biological therapy (monoclonal or polyclonal antibodies, vaccines, gene therapy, etc.) is also increasingly important^25^. Therefore, the innovative prognostic model constructed in this study may provide new insights for precision immunotherapy.

As an extension of cancer diagnosis, cancer subtype analysis can be considered a consensus clustering problem. This analysis is conducive to providing more accurate treatment for patients^26^. Consistency cluster analysis has been used to combine clinical research data with data from The Cancer Genome Atlas. The principle is to use the dataset to discover new subtypes and identify biological processes specific to different subtypes^27^. Currently, its application is not limited to genomics research; the consensus clustering analysis model based on CT radiomics is also gradually gaining popularity^28^. Therefore, we applied this method to CRC to identify significant differences among subgroups.

Interleukin 13 (IL-13) is an immune regulatory cytokine that is mainly secreted by Th2-like cells and natural killer T cells^29^. Among the CRC samples, IL-13 receptor overexpression was detected in more than 66% of the tumor samples. Moreover, in the immunohistochemistry analysis of stage I-III CRC patients, high IL-13 and IL-13R receptor expression was observed. 50% and 42%^30^. Saigusa studied 241 CRC patients and reported that serum IL-13 levels were significantly reduced in advanced patients and that low serum IL-13 levels were significantly correlated with poor prognosis^31^. Studies have reported that IL-13 overexpression promotes CRC liver metastasis by activating PI3K/AKT-related signaling pathways and that FAM120A in the IL-13/IL-13R signaling pathway is a key mediator of CRC invasion and liver metastasis^32^. IL-13R silencing inhibits the IL-13-induced proliferation of CRC cell lines through the downregulation of STAT6 activation^33^. These findings reveal the hub role of IL-13 in the occurrence and development of CRC, which provides a theoretical basis for targeting and immunotherapy in CRC patients. The tumor suppressor gene CDKN2A (P16) is a major target of carcinogenesis, with a frequency second only to that of the p53 tumor suppressor gene^34^. A recent study revealed that CDKN2A mRNA expression acts as a biomarker of clinically invasive meningiomas, with potential therapeutic significance^35^. A meta-analysis of CRC revealed a significant association between CDKN2A hypermethylation and lymphovascular invasion, lymph node metastasis, and proximal tumor location^36^. Another observational study revealed that the absence of the tumor suppressor CDKN2A was associated with the risk of CRC recurrence^37^. BEND3 is a four-member BEN domain-containing protein that binds to heterochromatin and functions as a transcription repressor^38^. The lncRNA XLOC_011677-miR-10b-5p-BEND3 potential exosomal ceRNA regulatory network may reveal regulatory pathways and diagnostic biomarkers related to the molecular biology^39^ of CRC. EXOC3L2 was previously considered one of the susceptibility genes for Alzheimer’s disease, and its study of cancer may become a new direction for future research^40^. CAPN9 is expressed mainly in the stomach and small intestine and is involved in the regulation of hypertension, heart disease and gastric mucosal defense. It also potentially plays a role in the upregulation of metalloproteinases in the structural changes of the artery wall^41,42^.

miR-585-3p inhibits the proliferation and migration of high-grade serous ovarian cancer cells by targeting CAPN9^43^. In our study, longevity-related genes were used for the first time to develop a prognostic model of CRC and played an important role in advancing tumor research.

In this study, we integrated data from a total of 689 colorectal cancer (CRC) patients obtained from the TCGA and GEO databases, employing bioinformatics approaches, including multi-omics analyses, to investigate the effects of longevity-associated genes on CRC in detail. The construction of a clinically relevant prognostic model offers new avenues for research aimed at potentially improving the prognosis of CRC patients. We also explored the utility of the risk score in predicting responses to immunotherapy and analyzed how varying risk scores influence the differential expression of immune cells within tumors. Despite these advancements, our study has several limitations. First, there is an overreliance on publicly available database data. Second, the model requires validation through large, multicenter, prospective clinical trials to confirm its applicability. Finally, this study lacks experimental validation, and there are no reliable experimental data to support more comprehensive investigations.

## 5 Conclusion

In this study, we constructed a prognostic model based on five longevity-associated genes, which also elucidated the key interactions between longevity-associated genes and CRC-specific immune cells. This innovative prognostic model provides new insights for precision immunotherapy, which can be used to fight this disease and improve the prognosis of patients, thus opening a new way for personalized treatment selection.

## Data Availability

N/A

## Abbreviations

TME: tumor microenvironment
CAR: chimeric antigen receptor
PCR: Polymerase Chain Reaction
GWAS: Genome-Wide Association Studies
PCA: principal component analysis
DEGs: differentially expressed genes
GO: Gene Ontology
KEGG: Kyoto Encyclopedia of Genes and Genomes
LASSO: minimum absolute shrinkage and selection operator
AUC: area under the curve
OS: overall survival
IL-13: Interleukin 13

## Author Contributions

All authors contributed to the study conception and design. Material preparation, data collection and analysis were performed by Guangtao Min, Hongpeng Wang. The first draft of the manuscript was written by Yichu Huang and all authors commented on previous versions of the manuscript. All authors read and approved the final manuscript.

## Acknowledgments

We would like to express our gratitude to the TCGA and GEO datebases studies for sharing the data. We also acknowledgments all persons who participated in the study.

## Ethical Statement

All data from publicly available TCGA and GEO databases, ethical approval is therefore not required.

## References

1 Sung, H. et al. Global Cancer Statistics 2020: GLOBOCAN Estimates of Incidence and Mortality Worldwide for 36 Cancers in 185 Countries. CA Cancer J Clin 71, 209–249, doi:10.3322/caac.21660 (2021).

2 Zheng, R. S. et al. [Cancer incidence and mortality in China, 2022]. Zhonghua Zhong Liu Za Zhi 46, 221–231, doi:10.3760/cma.j.cn112152-20240119-00035 (2024).

3 Müller, F. et al. CD19 CAR T-Cell Therapy in Autoimmune Disease - A Case Series with Follow-up. N Engl J Med 390, 687–700, doi:10.1056/NEJMoa2308917 (2024).

4 Wang, H. et al. Catalase-positive Staphylococcus epidermidis based cryo-millineedle platform facilitates the photo-immunotherapy against colorectal cancer via hypoxia improvement. J Colloid Interface Sci 676, 506–520, doi:10.1016/j.jcis.2024.07.145 (2024).

5 Liu, G. et al. A Review on Drug Delivery System for Tumor Therapy. Front Pharmacol 12, 735446, doi:10.3389/fphar.2021.735446 (2021).

6 Beekman, M. et al. Genome-wide association study (GWAS)-identified disease risk alleles do not compromise human longevity. Proc Natl Acad Sci U S A 107, 18046–18049, doi:10.1073/pnas.1003540107 (2010).

7 Vincze, O. et al. Cancer risk across mammals. Nature 601, 263–267, doi:10.1038/s41586-021-04224-5 (2022).

8 Gorbunova, V., Seluanov, A., Zhang, Z., Gladyshev, V. N. & Vijg, J. Comparative genetics of longevity and cancer: insights from long-lived rodents. Nat Rev Genet 15, 531–540, doi:10.1038/nrg3728 (2014).

9 Zhang, Y. et al. Risk-stratified screening and colorectal cancer incidence and mortality: A retrospective study from the Prostate, Lung, Colorectal, and Ovarian Cancer Screening Trial. Prev Med 187, 108117, doi:10.1016/j.ypmed.2024.108117 (2024).

10 Nikolich-Žugich, J. The twilight of immunity: emerging concepts in aging of the immune system. Nat Immunol 19, 10–19, doi:10.1038/s41590-017-0006-x (2018).

11 Roma-Rodrigues, C., Mendes, R., Baptista, P. V. & Fernandes, A. R. Targeting Tumor Microenvironment for Cancer Therapy. International Journal of Molecular Sciences 20, 840 (2019).

12 Kasprzak, A. The Role of Tumor Microenvironment Cells in Colorectal Cancer (CRC) Cachexia. Int J Mol Sci 22, doi:10.3390/ijms22041565 (2021).

13 Hissong, E., Crowe, E. P., Yantiss, R. K. & Chen, Y. T. Assessing colorectal cancer mismatch repair status in the modern era: a survey of current practices and re-evaluation of the role of microsatellite instability testing. Mod Pathol 31, 1756–1766, doi:10.1038/s41379-018-0094-7 (2018).

14 Deelen, J. et al. A meta-analysis of genome-wide association studies identifies multiple longevity genes. Nat Commun 10, 3669, doi:10.1038/s41467-019-11558-2 (2019).

15 Ritchie, M. E. et al. limma powers differential expression analyses for RNA-sequencing and microarray studies. Nucleic Acids Res 43, e47, doi:10.1093/nar/gkv007 (2015).

16 Mukhopadhyay, P. et al. Log-Rank Test vs MaxCombo and Difference in Restricted Mean Survival Time Tests for Comparing Survival Under Nonproportional Hazards in Immuno-oncology Trials: A Systematic Review and Meta-analysis. JAMA Oncol 8, 1294–1300, doi:10.1001/jamaoncol.2022.2666 (2022).

17 Mayakonda, A., Lin, D. C., Assenov, Y., Plass, C. & Koeffler, H. P. Maftools: efficient and comprehensive analysis of somatic variants in cancer. Genome Res 28, 1747–1756, doi:10.1101/gr.239244.118 (2018).

18 Kang, J. et al. LASSO-Based Machine Learning Algorithm for Prediction of Lymph Node Metastasis in T1 Colorectal Cancer. Cancer Res Treat 53, 773–783, doi:10.4143/crt.2020.974 (2021).

19 Pan, W., Huang, W., Zheng, J., Meng, Z. & Pan, X. Construction of a prognosis model of head and neck squamous cell carcinoma pyroptosis and an analysis of immuno-phenotyping based on bioinformatics. Transl Cancer Res 13, 299–316, doi:10.21037/tcr-23-922 (2024).

20 Chen, B., Khodadoust, M. S., Liu, C. L., Newman, A. M. & Alizadeh, A. A. Profiling Tumor Infiltrating Immune Cells with CIBERSORT. Methods Mol Biol 1711, 243–259, doi:10.1007/978-1-4939-7493-1_12 (2018).

21 Ruby, J. G. et al. Estimates of the Heritability of Human Longevity Are Substantially Inflated due to Assortative Mating. Genetics 210, 1109–1124, doi:10.1534/genetics.118.301613 (2018).

22 van den Berg, N., Beekman, M., Smith, K. R., Janssens, A. & Slagboom, P. E. Historical demography and longevity genetics: Back to the future. Ageing Res Rev 38, 28–39, doi:10.1016/j.arr.2017.06.005 (2017).

23 Tao, M. et al. Identification of a longevity gene through evolutionary rate covariation of insect mito-nuclear genomes. Nat Aging 4, 1076–1088, doi:10.1038/s43587-024-00641-z (2024).

24 Lu, B. et al. Evaluation of long-term benefits and cost-effectiveness of nation-wide colorectal cancer screening strategies in China in 2020-2060: a modelling analysis. Lancet Reg Health West Pac 51, 101172, doi:10.1016/j.lanwpc.2024.101172 (2024).

25 Duarte, D. & Vale, N. Combining repurposed drugs to treat colorectal cancer. Drug Discov Today 27, 165–184, doi:10.1016/j.drudis.2021.09.012 (2022).

26 Li, J., Xie, L., Xie, Y. & Wang, F. Bregmannian consensus clustering for cancer subtypes analysis. Comput Methods Programs Biomed 189, 105337, doi:10.1016/j.cmpb.2020.105337 (2020).

27 Manganaro, L. et al. Consensus clustering methodology to improve molecular stratification of non-small cell lung cancer. Sci Rep 13, 7759, doi:10.1038/s41598-023-33954-x (2023).

28 Jia, J., Liu, Z., Wang, F. & Bai, G. Consensus Clustering Analysis Based on Enhanced-CT Radiomic Features: Esophageal Squamous Cell Carcinoma patients’ 3-Year Progression-Free Survival. Acad Radiol 31, 2807–2817, doi:10.1016/j.acra.2023.12.025 (2024).

29 Hershey, G. K. IL-13 receptors and signaling pathways: an evolving web. J Allergy Clin Immunol 111, 677–690; quiz 691, doi:10.1067/mai.2003.1333 (2003).

30 Formentini, A. et al. Expression of interleukin-4 and interleukin-13 and their receptors in colorectal cancer. International Journal of Colorectal Disease 27, 1369–1376, doi:10.1007/s00384-012-1456-0 (2012).

31 Saigusa, S. et al. Low serum interleukin-13 levels correlate with poorer prognoses for colorectal cancer patients. Int Surg 99, 223–229, doi:10.9738/intsurg-d-13-00259.1 (2014).

32 Bartolomé, R. A. et al. IL13 Receptor α2 Signaling Requires a Scaffold Protein, FAM120A, to Activate the FAK and PI3K Pathways in Colon Cancer Metastasis. Cancer Res 75, 2434–2444, doi:10.1158/0008-5472.Can-14-3650 (2015).

33 Matsui, S. et al. Interleukin-13 and its signaling pathway is associated with obesity-related colorectal tumorigenesis. Cancer Sci 110, 2156–2165, doi:10.1111/cas.14066 (2019).

34 Liggett, W. H., Jr. & Sidransky, D. Role of the p16 tumor suppressor gene in cancer. J Clin Oncol 16, 1197–1206, doi:10.1200/jco.1998.16.3.1197 (1998).

35 Wang, J. Z. et al. Increased mRNA expression of CDKN2A is a transcriptomic marker of clinically aggressive meningiomas. Acta Neuropathol 146, 145–162, doi:10.1007/s00401-023-02571-3 (2023).

36 Xing, X. et al. The prognostic value of CDKN2A hypermethylation in colorectal cancer: a meta-analysis. Br J Cancer 108, 2542–2548, doi:10.1038/bjc.2013.251 (2013).

37 Berg, M. et al. Molecular subtypes in stage II-III colon cancer defined by genomic instability: early recurrence-risk associated with a high copy-number variation and loss of RUNX3 and CDKN2A. PLoS One 10, e0122391, doi:10.1371/journal.pone.0122391 (2015).

38 Kurniawan, F. et al. BEND3 safeguards pluripotency by repressing differentiation-associated genes. Proc Natl Acad Sci U S A 119, doi:10.1073/pnas.2107406119 (2022).

39 Zhao, Y., Song, X., Song, X. & Xie, L. Identification of Diagnostic Exosomal LncRNA-miRNA-mRNA Biomarkers in Colorectal Cancer Based on the ceRNA Network. Pathol Oncol Res 28, 1610493, doi:10.3389/pore.2022.1610493 (2022).

40 Seshadri, S. et al. Genome-wide analysis of genetic loci associated with Alzheimer disease. Jama 303, 1832–1840, doi:10.1001/jama.2010.574 (2010).

41 Logan, J. G., Yun, S., Bao, Y., Farber, E. & Farber, C. R. RNA-sequencing analysis of differential gene expression associated with arterial stiffness. Vascular 28, 655–663, doi:10.1177/1708538120922650 (2020).

42 Cui, H. X. et al. Expression and effect of Calpain9 gene genetic polymorphism on slaughter indicators and intramuscular fat content in chickens. Poult Sci 97, 3414–3420, doi:10.3382/ps/pey232 (2018).

43 Lu, X., Li, G., Liu, S., Wang, H. & Chen, B. MiR-585-3p suppresses tumor proliferation and migration by directly targeting CAPN9 in high grade serous ovarian cancer. J Ovarian Res 14, 90, doi:10.1186/s13048-021-00841-w (2021).

